# Dietary diversity and its correlates among pregnant adolescent girls in Ghana

**DOI:** 10.1101/2020.08.06.20169383

**Authors:** Linda A. Gyimah, Reginald A. Annan, Charles Apprey, Anthony K. Edusei, Linda Esi Aduku, Odeafo Asamoah-Boakye, Wisdom Azanu, Herman Lutterodt

**Affiliations:** Department of Biochemistry and Biotechnology, KNUST; Komfo Anokye Teaching Hospital, Kumasi, Ashanti Region; Department of Food Science and Technology, KNUST; Department of Community Health, School of Public Health, KNUST

**Keywords:** Dietary diversity, pregnant adolescents, eating behaviour, lived poverty index, household hunger scale

## Abstract

**Background:** Dietary diversity, a qualitative measure of dietary intake, which reflects the variety of foods consumed has been recommended to assuage nutritional problems related to insufficient micronutrients and food insecurity. To better understand the underlying factors for poor birth outcomes in Ghana, we assessed factors associated with dietary diversity among rural and urban pregnant adolescents in the Ashanti Region of Ghana.

**Methods:** As part of a larger longitudinal cohort of 416 pregnant adolescents, the FAO women’s dietary diversity index was used to determine dietary diversity score (DDS) of the participants from a 3-day repeated 24-hour dietary recall data. The household hunger scale (HHS) and lived poverty index (LPI) were used to determine hunger and socioeconomic status. Eating behavior and socio-demographic data were gathered using interviewer-administered questionnaires.

**Results:** The mean DDS for participants was 4.3 but 59.7% of participants were below the minimum DD. More rural (67.1%) than urban dwellers (53.9%) had inadequate DDS (p=0.008). Urban dwelling adolescents recorded higher odds for adequate DD (OR=1.8 CI =1.0-2.8, p=0.034) compared to the rural respondents. Other factors associated with higher odds for adequate DD are income (OR=12.1, p = 0.043, 95%CI= 1.1-136.8), moderate (OR=2.1, p = 0.019, 95%CI=1.1-3.8) and high LPI (OR=2.2, p=0.007, 95%CI=1.2-3.9), practicing food aversion (OR=1.6, p = 0.046, 95%CI= 1.0-2.4), and pica (OR 1.6, p =0.043, 95%CI= 1.0-2.5).

**Conclusions:** Low DD is common among pregnant adolescents in this study and associated with poverty, food insecurity, rural living, pica practice, and food aversions. Livelihood support for pregnant teenagers and nutrition education are recommended interventions to improve dietary quality and limit the consequences of poor dietary diversity.

## 1. INTRODUCTION

Pregnancy poses a significant risk for malnutrition (1) because extra nutrients are needed to support the growth of the developing foetus. For the adolescent, this risk is heightened due to the continuous psychological, social, and physical development of the pregnant girl (2). Food insecurity and malnutrition continue to be a challenge worldwide and developing countries are the most affected (3). To lessen the causes of malnutrition globally, the first two goals of the Sustainable Development Goals (SDGs) seek to reduce poverty and end all forms of malnutrition and hunger by 2030 (4). Accordingly, one major contributor to maternal malnutrition in developing countries is inadequate dietary intake mainly due to inadequate quantity, quality and diversified diets, poverty, high-intensity agricultural labor, and high fertility rate (5). Dietary diversity is one of the recommended strategies to improve maternal diet and prevent malnutrition, through increased daily intake of the various food groups in one’s diet (6).

Dietary diversity (DD) has also been used as a representation for nutrient adequacy and quality of dietary intake and particularly, the probability of micronutrient adequacy in a diet (7). It is a qualitative assessment of food intake that reflects an individual’s access to a variety of foods. The Women’s Dietary Diversity Score (WDDS), which involves the classification of a previous day’s meals into nine food groups is used to measure the dietary diversity of women of reproductive age (7). Studies have reported significant associations between dietary diversity and nutrient adequacy (8-10), dietary diversity and socioeconomic status (11) as well as dietary diversity and household food security (12).

Household hunger scale (HHS) is used to assess food deprivation in a population and together with other tools like DD and income, HHS can be used to measure some aspects of food security (13). Lived poverty is an index that measures the frequency with which people experience shortages of basic necessities during the year (14). LPI measures a portion of the concept of poverty that is not well assessed by other measures. Several authors have investigated that people living in poverty are not exposed to resources which improve health such as food market, a supply of nutritious food, water quality, recreational facilities, housing conditions, employment, education and access to medical care (15-17).

During pregnancy, eating behaviors change. Food cravings and aversions are indicators of physiological stress during pregnancy (18,19) as a result of changes in metabolic states and hormone levels (20). Pregnant women usually crave sweet, salty, fatty, or spicy foods which are generally considered unhealthy (21) and avert certain foods that can limit the intake of a variety of dishes. Studies on food craving and aversions indicate that pregnant women usually crave energy-dense foods (22) and avert nutritious diets such as cereals, meat, fish (18,23) and fruits like mangoes and pineapples (24). Pica, another form of craving involves the consumption of non-food substances like clay, soap, ashes, paint, paper. Pica has been of public interest due to its high prevalence in pregnant women. It is useful in protecting against pathogens and toxins, quelling nausea, vomiting, diarrhea (25). It may also be harmful as it reduces the bioavailability of beneficial nutrients and introducing toxic substances (26).

Adolescent pregnancy in Ghana has been linked to socio-economic status, ignorance, and their inability to access information and services related to reproductive health (27), but dietary diversity and factors associated with it among pregnant adolescents in Ghana have not been studied. In Ghana, about 14.4% of adolescent girls aged between 15 and 19 years have begun childbearing and about 12% have a live birth (28). Since a diverse diet indicates a healthier diet, it suggests that good dietary diversity can influence good pregnancy outcomes. Therefore, understanding DD issues among pregnant adolescents is a step towards addressing malnutrition and its effects on pregnancy outcomes. This study assessed the dietary diversity and its correlates associated with pregnant adolescents in Ashanti Ghana.

## 2. METHOD

### STUDY SETTING

The Ashanti Region is located in the southern sector of Ghana occupying about 10% of the total land area of Ghana. In terms of population, it is the most populated region in Ghana, with an estimated population of 5,792,187, accounting for 19% of Ghana’s total population (29). The region is divided into 31 districts with Kumasi Metropolis being the most populated and the second largest city in Ghana. Each district has a district hospital and several Community-based Health Planning Services (CHPS) compounds/health centres depending on the size of the district (30). Due to its position in the middle belt of Ghana, the Ashanti region was chosen for this study and also because the majority of maternal and child health studies have been conducted in both the Northern and Southern sectors of Ghana (31,32). A rural area is a community with less than 5000 population while all others are considered urban (30). By this classification, the Ashanti region is about 61% urban.

### STUDY DESIGN

This study is part of a larger longitudinal study among pregnant adolescents (aged 13 to 19 years old) to assess the impact of nutrition and birth outcomes. The study involves 416 pregnant adolescents with gestational age up to 32 gestational weeks, recruited from May to August 2018. The study was conducted in 29 communities in three urban districts (Kumasi Metropolis, Asante Akim Central and Ejisu Juaben) and five rural districts (Bosumtwi, Asante Akim South and North and Ahafo Ano North and South) in Ashanti Region, Ghana.

### SAMPLING

The sample size of 420 was calculated based on a low birth weight prevalence of 23% reported in a previous study by (33) and a marginal error of 5%. However, 416 pregnant adolescents were recruited within the period of data collection. Participants were recruited from hospitals and health centres during maternity care visits. Most of these hospitals have maternity clinics days for only pregnant adolescents. On these dates, researchers visited the facilities, and any pregnant adolescent within the required age group who gave consent was recruited for the study. For health centres in rural areas, announcements were made at community information centres to invite pregnant adolescents to the health centres on specific dates. These announcements were necessary as the adolescents may not normally visit the health centres due to stigma.

### DATA COLLECTION

A standardized questionnaire were used to collect data on the socio-demographic variables, dietary diversity, eating behaviours (pica, food aversion, food craving), food deprivation (household hunger scale) and availability of basic necessities (lived poverty index) of the pregnant adolescents. Data on age and parity were verified from their National Health Insurance Identification cards and maternal health record book. The questionnaire was pretested and validated to ensure appropriate responses from participants. The dependent variable for the study was dietary diversity whilst community type, age, parity, occupation, income, level of education, marital status, pica, food aversion, food craving, lived poverty index and household hunger scale.

### ASSESSMENT OF DIETARY DIVERSITY AND EATING BEHAVIOUR

Dietary diversity was assessed using the repeated 24-hour dietary recall method which was taken three times (two weekdays and one weekend). Participants were asked to recall food taken during the three previous days before the interview day. The FAO’s Women’s Dietary Diversity (WDDS) was used to determine maternal dietary diversity (34). The WDDS comprises of ten (10) food groups, namely; grains, white roots and tubers, plantain; pulses (beans, pea, lentils); nuts and seeds; dairy; meat, poultry, and fish; eggs; dark green leafy vegetables; other vitamin A rich fruits and vegetables; other vegetables; other fruits (34). The first 24-hour recall from each participant were used to score the dietary diversity using the 10 food groups. The WDDS was then categorised as low (≤ 3), medium (4 – 5 food groups), or high (≥ 6 food groups) according to FAO’s guidelines (7). The WDDS was further categorized into adequate (5 – 9) and inadequate (0 – 4) dietary diversity scores to help with statistical association for the population. A questionnaire was used to obtain data on food cravings, pica practices, and food aversions.

### ASSESSMENT OF FOOD AVAILABILITY AND POVERTY

The HHS was assessed through interviews by asking participants questions related to food availability and deprivation over the past month. The responses to these questions are coded and scored which ranges between 0 – 6. Scores between < 2 means little or no hunger, 2 – 3 means moderate hunger, and 4 – 6 means severe hunger (13). The Lived Poverty Index (LPI) is the sum of responses given to a set of questions about the availability of food, water, cash income, medical care, and cooking fuel over the past year. The responses are scored ranging along a five-point scale from 0 (which can be thought of as no lived poverty) to 4 which would reflect a constant absence of all basic necessities (14). The averages of the scores were then categorized into low (0 – 0.05), low-moderate (0.51-1.0), high-moderate (1.01-1.5) and high (> 1.5) LPI.

### ETHICAL CLEARANCE

Ethical clearance for the study (Reference: CHPRE/ AP/236/18) was obtained from Committee on Human Research, Publications and Ethics of the Kwame Nkrumah University of Science and Technology, Kumasi, Ghana (CHRPE/KNUST) and personal or parental consent were obtained from participants before the beginning of the study.

### DATA ANALYSIS

Data collected were first entered into Microsoft Excel 2019 and thoroughly cleaned to eliminate errors in the data. Then, data was imported into Statistical Package for Social Sciences version 25 (SPSS IBM Inc Chicago, USA) for statistical analysis. sociodemographic characteristics, eating behaviours, hunger, and poverty status were used as independent variables while dietary diversity was used as the main outcome variable (dependent variable). Descriptive statistics was done and reported as relative frequencies for sociodemographic characteristics, eating behaviours, hunger and poverty status, and dietary diversity. A test of normality using Kolmogorov-Smirnov test was done to ascertain whether the continuous variables were normally distributed. A chi-square (Fisher’s exact test) cross-tabulation was performed to compare frequencies of sociodemographic characteristics, eating behaviours, hunger, and poverty status by community type and dietary diversity. Independent t-test and one-way ANOVA were used for parametric mean comparisons of the continuous variables. A non-parametric test (Mann Whitney ‘U’ test and Kruskal Wallis test) was performed to compare mean differences between age, HHS, LPI, and DDS. A bivariate correlation was used to determine the association between age, HHS, LPI, and DDS. A binary logistic regression analysis was performed to determine predictors of dietary diversity, and these are reported as odds ratios, to explain the combined effect size of the independent variables. All tests were 2-tailed, and differences were considered statistically significant at p < 0.05.

## 3. RESULTS

Sociodemographic characteristics of participants are presented in Table 1. Majority of the participants were unmarried (76.0%), unemployed (71.6%), lived on no income (74.5%), had only completed basic education (61.3%) and were between the ages 16 and 19 years (92.3%). More rural than urban participants were younger teenagers (8.1% versus 7.4%, p = 0.853) and had more parity (26.0% versus 22.6%, p = 0.485). More urban (73.7%) than rural (68.8%) pregnant adolescents were unemployed (p = 0.321). More urban (25.1%) than rural (14.5%) pregnant adolescents completed secondary education (p = 0.018).

**Table 1.** Distribution of Socio-Demographic Characteristics of Participants.

| CHARACTERISTICS | TOTAL (%) | RURAL (%) | URBAN (%) | P VALUE |
| --- | --- | --- | --- | --- |
| **Age group (Years)** |  |  |  |  |
| 13 - 15 | 32 (7.7) | 14 (8.1) | 18 (7.4) | 0.853<sup>¥</sup> |
| 16 - 19 | 384 (92.3) | 159 (91.9) | 225 (92.6) |  |
| **Mean age (years)** | **17.5 ± 1.4** |  |  |  |
| **Total, n (%)** | **416 (100)** | **173 (41.6)** | **243 (58.4)** |  |
| **Marital Status** |  |  |  |  |
| Single | 316 (76.0) | 134 (77.5) | 182 (74.9) | 0.563<sup>¥</sup> |
| Married | 100 (24.0) | 39 (22.5) | 61 (25.1) |  |
| **Occupation** |  |  |  |  |
| Unemployed | 298 (71.6) | 119 (68.8) | 179 (73.7) | 0.321<sup>¥</sup> |
| Employed | 118 (28.4) | 54 (31.2) | 64 (26.3) |  |
| **Parity** |  |  |  |  |
| 1 | 316 (76.0) | 128 (74.0) | 188 (77.4) | 0.485<sup>¥</sup> |
| >1 | 100 (24.0) | 45 (26.0) | 55 (22.6) |  |
| **Education** |  |  |  |  |
| None | 19 (4.6) | 8 (4.6) | 11 (4.5) | **0.018<sup>‡</sup>** |
| Primary | 56 (13.5) | 31 (17.9) | 25 (10.3) |  |
| JHS | 255 (61.3) | 109 (63) | 146 (60.1) |  |
| SHS | 86 (20.7) | 25 (14.5) | 61 (25.1) |  |
| **Income (C)** |  |  |  |  |
| No Income | 310 (74.5) | 135 (78.0) | 175 (72.0) | 0.173<sup>¥</sup> |
| Below 100 | 41 (9.9) | 19 (11.0) | 22 (9.1) |  |
| Between 500 | 61 (14.7) | 18 (10.4) | 43 (17.7) |  |
| More than 500 | 4 (1) | 1 (0.6) | 3 (1.2) |  |

Data are reported as frequency (percentage), JHS- Junior High School, SHS- Senior High School, <sup>‡</sup> Chi-Square p value, some cells were less than 5 for chi-square, <sup>¥</sup> Fisher's exact test p value. bold values are significant at $p < 0.05$ .

Table 2 presents household Hunger, Lived Poverty Index, Dietary Diversity, and Eating behaviour of Participants. More than a fifth of the participants (27.1%) were food-deprived, while 36.5%, fell within the high poverty category. More than a third (38.0%) of the participants practiced pica, 64.2%, did crave for food and non-food substances, and 44.5%, had food aversion during pregnancy. Also, more than half of the participants (59.4%) had inadequate dietary diversity. A higher proportion of rural than urban participants were deprived of food (27.2% versus 1.2%, p<0.001) and lived in abject poverty (51.4% versus 25.9%, p<0.001). Although more rural participants practiced food craving, pica was practiced higher among urban dwellers than rural dwellers.

**Table 2.** Household Hunger, Lived Poverty Index, Dietary Diversity and Eating Behaviours of Participants.

| VARIABLES | TOTAL (%) | RURAL (%) | URBAN (%) | P VALUE |
| --- | --- | --- | --- | --- |
| **Household Hunger Scale** |  |  |  | **<0.001<sup>¥</sup>** |
| No Hunger | 303 (72.8) | 88 (50.9) | 215 (88.5) |  |
| Moderate Hunger | 63 (15.1) | 38 (22.0) | 25 (10.3) |  |
| Severe Hunger | 50 (12.0) | 47 (27.2) | 3 (1.2) |  |
| **Lived Poverty Index** |  |  |  | **<0.001<sup>‡</sup>** |
| Low Moderate | 194 (45.2) | 50 (28.9) | 138 (56.8) |  |
| High Moderate | 75 (18.3) | 34 (19.7) | 42 (17.3) |  |
| High | 152 (36.5) | 89 (51.4) | 59 (25.9) |  |
| **Food Aversion** |  |  |  | **0.046<sup>¥</sup>** |
| Yes | 185 (44.5) | 87 (50.3) | 98 (40.3) |  |
| No | 231 (55.5) | 86 (49.7) | 145 (59.7) |  |
| **Food Craving** |  |  |  | **0.078<sup>¥</sup>** |
| Yes | 267 (64.2) | 120 (69.4) | 147 (60.5) |  |
| No | 149 (35.8) | 53 (30.6) | 96 (39.5) |  |
| **PICA Practice** |  |  |  | **0.032<sup>¥</sup>** |
| Yes | 160 (38.5) | 56 (32.4) | 104 (42.8) |  |
| No | 256 (61.5) | 117 (67.6) | 139 (57.2) |  |
| **Women's Dietary Diversity** |  |  |  | **0.659<sup>‡</sup>** |
| Low | 108 (26.0) | 48 (27.7) | 60 (24.7) |  |
| Medium | 285 (68.5) | 117 (67.6) | 168 (69.1) |  |
| High | 23 (5.5) | 8 (4.6) | 15 (6.2) |  |
| **Dietary Diversity** |  |  |  | **0.008<sup>¥</sup>** |
| Inadequate | 247 (59.4) | 116 (67.1) | 131 (53.9) |  |
| Adequate | 169 (40.6) | 57 (32.9) | 112 (46.1) |  |

Categorical data are presented as frequency (percentage), <sup>‡</sup> Chi-Square p value, <sup>¥</sup> Fisher's exact test p value. bold values are significant at $p < 0.05$ .

Figure 1 indicates the percentage of participants who went through pregnancy year without basic necessities. Overall, about 43% and 32% of the participants reported going without income and food at least once or twice in the past year respectively. Two in ten (23%) participants reported having shortages in medical care at least once or twice in the past year whilst approximately 7% of the participants went without water.

**Fig. 1.**
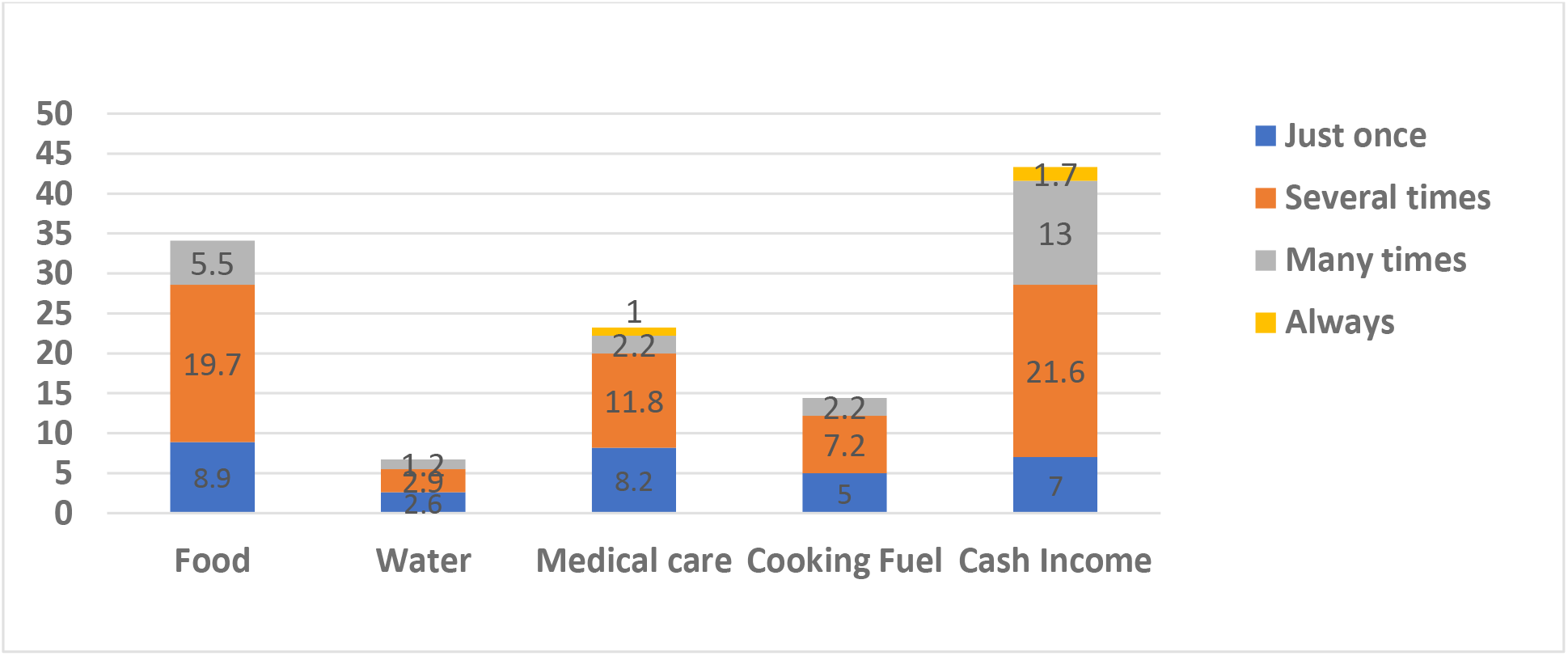
Percentage of Participants who went without Basic Necessities.

Table 3 presents proportions of sociodemographic characteristics who had adequate and inadequate dietary diversity. A higher proportion of the rural (67.1%) than urban (53.9%) pregnant adolescents had inadequate dietary diversity (p = 0.008). Participants who had lower educational achievement (that is, completed primary school) presented the largest proportion (67%) of those with inadequate dietary diversity. Dietary Diversity did not vary by age, marital status, parity, occupation, and income status of the pregnant girls.

**Table 3.** RELATIONSHIP BETWEEN DIETARY DIVERSITY AND SOCIO-DEMOGRAPHIC CHARACTERISTICS.

| VARIABLES | DIETARY DIVERSITY (DD) |  | X<sup>2</sup>, P VALUE | RR (95%CI) FOR INADEQUATE DD |
| --- | --- | --- | --- | --- |
|  | INADEQUATE | ADEQUATE |  |  |
| Community Type |  |  |  |  |
| Rural | 116 (67.1) | 57 (32.9) | 7.236, 0.008<sup>¥</sup> | 1.2(1.1-1.5) |
| Urban | 131 (53.9) | 112 (46.1) |  | 1.0 |
| Age group (Years) |  |  |  |  |
| 13 - 15 | 15 (46.9) | 17 (53.1) | 2.246, 0.139<sup>¥</sup> | 0.8(0.5-1.1) |
| 16 - 19 | 232 (60.4) | 152 (39.6) |  | 1.0 |
| Marital Status |  |  |  |  |
| Single | 192 (60.8) | 124 (39.2) | 1.045, 0.350<sup>¥</sup> | 1.1(0.9-1.3) |
| Married | 55 (55) | 45 (45) |  | 1.0 |
| Occupation |  |  |  |  |
| Unemployed | 168 (56.4) | 130 (43.6) | 3.918, 0.060<sup>¥</sup> | 0.8(0.7-1.0) |
| Employed | 79 (66.9) | 39 (33.1) |  | 1.0 |
| Parity |  |  |  |  |
| 1 | 184 (58.3) | 132 (41.8) | 0.717, 0.416<sup>¥</sup> | 0.9(0.7-1.1) |
| >1 | 63 (63.0) | 37 (37.0) |  | 1.0 |
| Education |  |  |  |  |
| None | 9 (47.4) | 10 (52.6) | 12.174, 0.007<sup>‡</sup> |  |
| Primary | 43 (76.8) | 13 (23.2) |  |  |
| JHS | 153 (60.0) | 102 (40.0) |  |  |
| SHS | 42 (48.8) | 44 (51.2) |  |  |
| Income |  |  |  |  |
| No Income | 192 (61.9) | 118 (38.1) | 6.387, 0.094<sup>¥</sup> |  |
| Below 100 | 25 (61.0) | 16 (39.0) |  |  |
| Between 100-500 | 29 (47.5) | 32 (52.5) |  |  |
| More than 500 | 1 (25.0) | 3 (75.0) |  |  |

Data are presented as frequency (percentage), <sup>‡</sup> Chi-Square, <sup>¥</sup> Fisher's exact test, RR- Relative risk, Bold values are significant at $p < 0.05$ .

Table 4 presents the relationship between dietary diversity and household hunger scale, lived poverty, and eating behaviour. HHS, LPI, and eating behavior did not vary significantly by dietary diversity (p>0.05). However, a large proportion of participants with low moderate LPI (61.7%, p=0.607) and severe hunger (68.0%, p=0.197) presented inadequate dietary diversity compared to the others. More participants who were practicing pica (62.5%), food aversion (63.8%), and food craving (57.5) had inadequate dietary diversity compared to those who did not (p>0.05).

**Table 4.** RELATIONSHIP BETWEEN DIETARY DIVERSITY AND HOUSEHOLD HUNGER SCALE, LIVED POVERTY AND EATING BEHAVIOURS.

| VARIABLES | DIETARY DIVERSITY |  | P VALUE | RR (95%CI) FOR INADEQUATE DD |
| --- | --- | --- | --- | --- |
|  | INADEQUATE | ADEQUATE |  |  |
| **Household Hunger Scale** |  |  |  |  |
| No Hunger | 172 (56.8) | 131 (43.2) | 3.247, 0.197<sup>‡</sup> |  |
| Moderate Hunger | 41 (65.1) | 22 (34.9) |  |  |
| Severe Hunger | 34 (68.0) | 16 (32.0) |  |  |
| **Low Poverty Index** |  |  |  |  |
| Low Moderate | 116 (61.7) | 72 (38.3) | 1.002, 0.607<sup>‡</sup> |  |
| High Moderate | 42 (55.3) | 34 (44.7) |  |  |
| High | 89 (58.6) | 63 (41.4) |  |  |
| **Food Aversion** |  |  |  |  |
| Yes | 118 (63.8) | 67 (36.2) | 2.685, 0.109<sup>¥</sup> | 1.1(1.0-1.3) |
| No | 129 (55.8) | 169 (44.2) |  | 1.0 |
| **Food Craving** |  |  |  |  |
| Yes | 154 (57.7) | 113 (42.3) | 0.890, 0.351<sup>¥</sup> | 0.9(0.8-1.1) |
| No | 93 (62.4) | 56 (37.6) |  | 1.0 |
| **Pica Practice** |  |  |  |  |
| Yes | 100 (62.5) | 60 (37.5) | 1.053, 0.356<sup>¥</sup> | 1.1(0.9-1.3) |
| No | 147 (57.4) | 169 (42.6) |  | 1.0 |

Data are presented as frequency (percentage), <sup>‡</sup> Chi-Square, <sup>¥</sup> Fisher's exact test, RR- Relative risk, DD- Diet diversity, P value is significant at $p<0.05$ .

Mean comparisons and correlation between age, household hunger scale, lived poverty and dietary diversity is displayed in Table 5. The total mean DDS for the population was 4.3 ± 0.060. The means DDS of participants who fell within no hunger (4.4 ± 1.2) was higher than those within moderate (4.1 ± 1.1) and severe hunger (4.0 ± 1.2) (p = 0.043). Participants within the ages 13 – 15 years presented a higher mean DDS (4.7 ± 1.2) compared to those aged 16 – 19 years (4.3 ± 1.2) (p = 0.052). The means of DDS did not vary by lived poverty index (p = 0.463). A weak, negative association existed between HHS and dietary diversity (r = −0.12, p = 0.018).

**Table 5.**
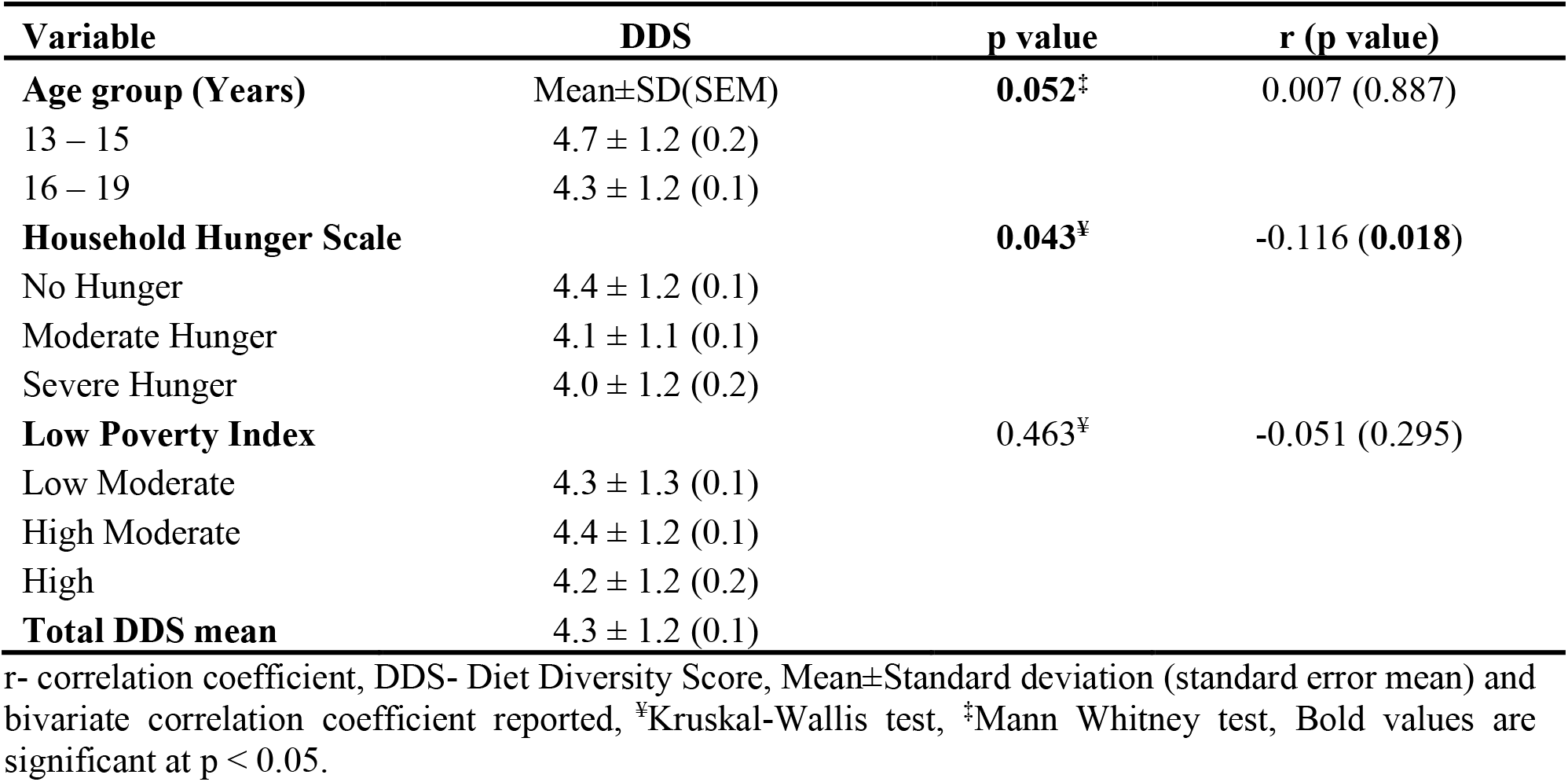
MEAN COMPARISON AND CORRELATION BETWEEN AGE, HOUSEHOLD.

| Variable | DDS | p value | r (p value) |
| --- | --- | --- | --- |
| **Age group (Years)** | Mean±SD(SEM) | **0.052<sup>‡</sup>** | 0.007 (0.887) |
| 13 – 15 | 4.7 ± 1.2 (0.2) |  |  |
| 16 – 19 | 4.3 ± 1.2 (0.1) |  |  |
| **Household Hunger Scale** |  | **0.043<sup>¥</sup>** | **-0.116 (0.018)** |
| No Hunger | 4.4 ± 1.2 (0.1) |  |  |
| Moderate Hunger | 4.1 ± 1.1 (0.1) |  |  |
| Severe Hunger | 4.0 ± 1.2 (0.2) |  |  |
| **Low Poverty Index** |  | 0.463<sup>¥</sup> | -0.051 (0.295) |
| Low Moderate | 4.3 ± 1.3 (0.1) |  |  |
| High Moderate | 4.4 ± 1.2 (0.1) |  |  |
| High | 4.2 ± 1.2 (0.2) |  |  |
| **Total DDS mean** | 4.3 ± 1.2 (0.1) |  |  |

r- correlation coefficient, DDS- Diet Diversity Score, Mean±Standard deviation (standard error mean) and bivariate correlation coefficient reported, <sup>¥</sup>Kruskal-Wallis test, <sup>‡</sup>Mann Whitney test, Bold values are significant at $p < 0.05$ .

The predictors of adequate dietary diversity of the pregnant adolescent are presented in Table 6. All the variables used were significantly associated with adequate DDS (≥5 food groups) except for HHS and parity. Urban dwellers (OR= 1.7, p= 0.034, 95%CI= 1.0-2.8) had higher odds for adequate DDS than rural. Pregnant adolescents who earned between 100 to 500 Ghana cedis (OR= 2.9, p = 0.001, 95%CI= 1.5-5.4), those who earned more than 500 Ghana cedis (OR=12.1, p = 0.043, 95%CI= 1.1-136.8) had higher odds for adequate DDS than non-earners of income. Pregnant adolescents with moderate lived poverty (OR= 2.1, p= 0.019, 95% CI 1.1-3.8), high lived poverty (OR=2.2, p= 0.007, 95%CI= 1.2-3.9), had lower odds for adequate DDS compared with those with no poverty. Those who averted food (OR=1.6, p= 0.046, 95%CI= 1.0-2.4) and those who were not practicing pica (OR=1.6, p= 0.043, 95%CI= 1.0-2.5) had higher odds for adequate dietary diversity than those who did not. Older adolescents (16 – 19 years bracket) (OR= 0.5, p= 0.061, 95%CI= 0.2-1.0), those with primary education (OR= 0.2, p= 0.010, 95% CI 0.1-0.7), those who did not practice food craving (OR= 0.6, p= 0.026, 95%CI= 0.4-0.9) and employed girls (OR= 0.5, p= 0.021, 95%CI= 0.3-0.9) had lower odds for adequate dietary diversity.

**Table 6.**
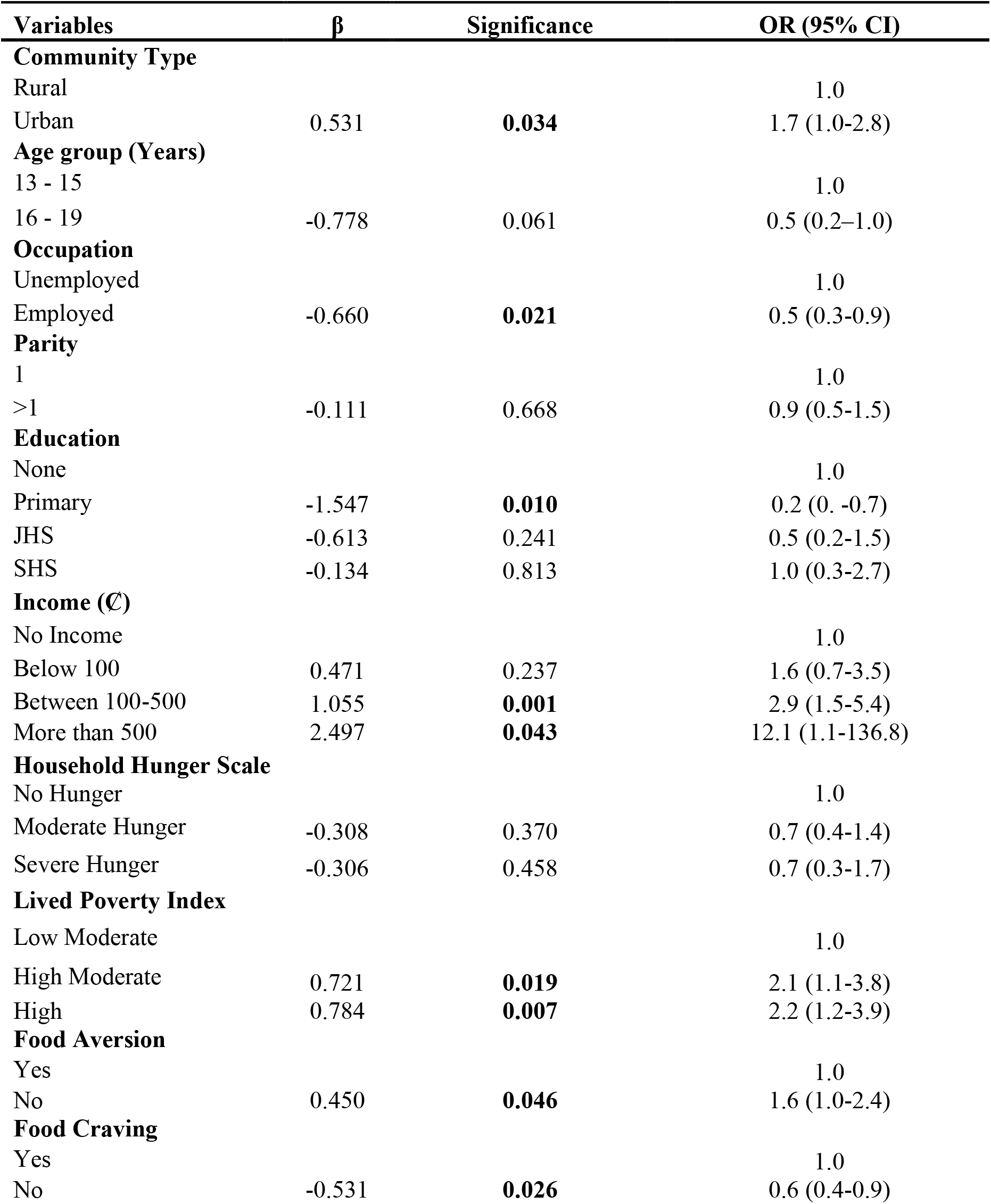
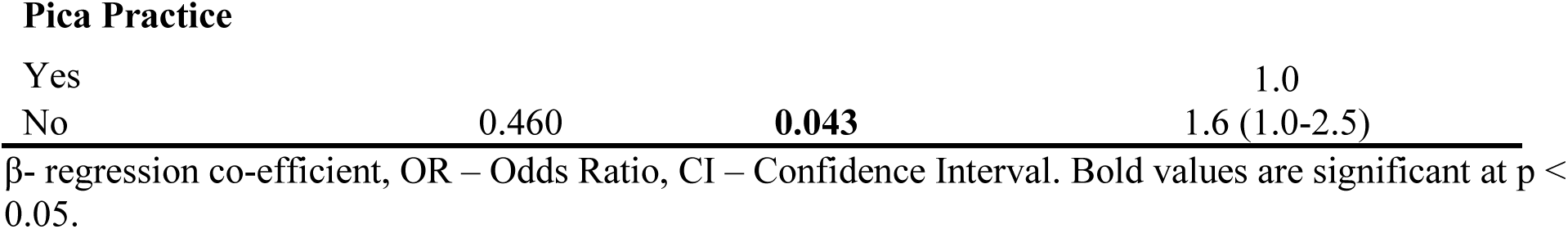
PREDICTORS OF ADEQUATE DIETARY DIVERSITY AMONG PREGNANT ADOLESCENTS.

| **Variables** | **β** | **Significance** | **OR (95% CI)** |
| --- | --- | --- | --- |
| **Community Type** |  |  |  |
| Rural |  |  | 1.0 |
| Urban | 0.531 | **0.034** | 1.7 (1.0-2.8) |
| **Age group (Years)** |  |  |  |
| 13 - 15 |  |  | 1.0 |
| 16 - 19 | -0.778 | 0.061 | 0.5 (0.2–1.0) |
| **Occupation** |  |  |  |
| Unemployed |  |  | 1.0 |
| Employed | -0.660 | **0.021** | 0.5 (0.3-0.9) |
| **Parity** |  |  |  |
| 1 |  |  | 1.0 |
| >1 | -0.111 | 0.668 | 0.9 (0.5-1.5) |
| **Education** |  |  |  |
| None |  |  | 1.0 |
| Primary | -1.547 | **0.010** | 0.2 (0. -0.7) |
| JHS | -0.613 | 0.241 | 0.5 (0.2-1.5) |
| SHS | -0.134 | 0.813 | 1.0 (0.3-2.7) |
| **Income (C)** |  |  |  |
| No Income |  |  | 1.0 |
| Below 100 | 0.471 | 0.237 | 1.6 (0.7-3.5) |
| Between 100-500 | 1.055 | **0.001** | 2.9 (1.5-5.4) |
| More than 500 | 2.497 | **0.043** | 12.1 (1.1-136.8) |
| **Household Hunger Scale** |  |  |  |
| No Hunger |  |  | 1.0 |
| Moderate Hunger | -0.308 | 0.370 | 0.7 (0.4-1.4) |
| Severe Hunger | -0.306 | 0.458 | 0.7 (0.3-1.7) |
| **Lived Poverty Index** |  |  |  |
| Low Moderate |  |  | 1.0 |
| High Moderate | 0.721 | **0.019** | 2.1 (1.1-3.8) |
| High | 0.784 | **0.007** | 2.2 (1.2-3.9) |
| **Food Aversion** |  |  |  |
| Yes |  |  | 1.0 |
| No | 0.450 | **0.046** | 1.6 (1.0-2.4) |
| **Food Craving** |  |  |  |
| Yes |  |  | 1.0 |
| No | -0.531 | **0.026** | 0.6 (0.4-0.9) |
| Pica Practice |  |  |  |
| Yes |  |  | 1.0 |
| No | 0.460 | **0.043** | 1.6 (1.0-2.5) |

$\beta$ - regression co-efficient, OR – Odds Ratio, CI – Confidence Interval. Bold values are significant at $p < 0.05$ .

## 4.0 DISCUSSION

This study sought to assess dietary diversity and its association with socio-demographic factors among pregnant adolescents in rural and urban districts of Ghana. Slightly more adolescents were recruited from the urban areas than the rural but this does not reflect the prevalence of teenage pregnancy in these areas as recruitment was limited to study period used in collecting data in these areas. The mean age (17.5 years) for the study population was similar to the reported mean age of pregnant adolescents in two rural districts in Ashanti region, Ghana (35). The minimum age of the participants was 13 years and about a quarter of the population had between 2 to 4 pregnancies at recruitment. These suggest that teenage pregnancy is starting at a much younger age in these districts and it explains why some teenagers had been pregnant several times. It also suggests early, unprotected sexual activities and unplanned pregnancies are common in these districts. The fact that less than 24% of these girls were married suggests that these girls are not being forced into marriage or sexual activities. Rather, most of them are likely to be school dropouts. This suggestion is corroborated by the level of education of the participants which found only 20.7% of the girls had completed secondary education during the study period. Since most girls would have finished basic education by ages 14-16 years, it could be that teenage pregnancy is driving low educational attainment. Therefore, these girls need social protection policies to empower them to focus on their education. It is also worth noting that just a quarter of these girls received income but were at the same time not married, meaning the care and support for their pregnancies go back to their parents, and can be definitely problematic. In our study, more of the pregnant teenagers from the rural areas were engaged in farming activities and this explains the higher proportion being employed as well as the higher proportion of rural pregnant adolescents receiving income compared to the urban group.

The mean DDS of our study participants was 4.3 representing a medium DDS but the urban girls were better off. More than half of the participants (59.4%) had inadequate intake or consumed less than 5 food groups in the last 3 days. Again, this situation is slightly better for the urban girls. The Ghana Micronutrient Survey in 2017 recorded a mean DDS of 4.40 among pregnant women of all ages (28). This finding is similar with the adolescent girls in our study, suggesting that DDS for adult pregnant women and adolescents in Ghana are similar. A study in the Northern part of Ghana (36) and Ashanti, Ghana (37) reported a mean DDS of 3.95 and 3.81 respectively which are slightly lower than our study. Studies in Bangladesh (38) and Kenya (39), respectively have reported higher WDDS of 5.1 and 5.6 among pregnant adolescents. Our study, the others in Ghana, and others compared outside Ghana, all suggested dietary diversity among pregnant women in general is inadequate. For the adolescents, the extra nutrient needs for their growth as well as that of the foetus make these inadequacies much more worrying. Inadequate DDS also depicts that these pregnant adolescents are likely to have nutrients deficiencies, especially micronutrients, which could have poor prognosis on birth outcome.

Socio-demographic factors have been showed to determine dietary intake in many populations (40-42). In this study, it is apparent that these variables drive the quality of dietary intake. Our findings showed that although more rural girls were employed and earned income, urban pregnant adolescents had better DDS (p = 0.008). A possible explanation is that improve access to quality diet are beyond income and employment but also availability that the food environment offers. The girls from the rural areas are likely to consume more monotonous meals from energy-dense foods (mainly starchy foods) they might have cultivated which is of a limited variety, compared to the urban girls where more varied food would be available to buy. In contrast, other studies (43,44) have reported that rural households with home garden and/or livestock rearing often have higher DDS. Therefore, interventions that promote rural household’s home gardening and poultry are recommended to boost availability and consumption of diverse food. It is also not apparent if certain socio-cultural factors and food taboos contribute to the low DDS among the rural girls as have been showed in this study (45), but this is worth investigating. Our findings strongly show that socio-demographic circumstances predicted DD of the pregnant adolescents. Residing in urban areas increased odds for a diverse diet. In contrast to what we found, (46)found no significant difference among rural and urban residents (P = 0.067). This could be due to improved access to diverse food generally in urban areas and agrees with (6,47) that poor communities were at higher risk of having low DDS. The income level of the adolescents also significantly increased the odds of having an adequate DDS by 2-12 times compared with not having income. Similarly, in Ethiopia, (6) reported a three times increased odds for adequate DDS of participants who received income. A significant association between adequate dietary diversity and education level of pregnant women has been established in several studies (12,38,48). In our findings, adolescents who have completed primary education presented reduced odds of having adequate dietary diversity. Though not significant, adolescents between 16-19 years also had lower odds of having adequate dietary diversity. This may suggest adequate dietary diversity reduced with age, evident in the 0.4 increase in the mean WDDS of younger girls (13-15 years).

The HHS assessment found that more than a quarter of the participants experienced hunger at least once or twice in a month. A larger proportion of this percentage were from the rural participants with a corresponding greater inadequate DD as compared to their urban folks. The proportion of participants experiencing hunger in the rural areas (49%) might be explained by the level of poverty (70%), since a greater proportion of the rural participants (27%) who experienced severe hunger, also showed high LPI. The mean LPI score for the study was 1.48, suggesting that the participants lived without each of the basic necessities once or twice over the past year before the study. The urban girls were again better. In a study to compare poverty status with total intake of micronutrients in USA, (49) found out that people living in poverty presented higher rates of inadequate nutrient intake. This also accords previous studies that age (50) and place of residence (51) are significant determinants of poverty in Kenya and Rwanda respectively. This suggests that pregnant adolescents living in poverty and deprived of food might be at risk of inadequate dietary intake. Those who were experiencing high lived poverty presented increased odds of having adequate dietary diversity twice. This relationship may partly be explained by the fact that those who have a high lived poverty index consume diverse diets regardless their limited budget. Thus, consuming a diverse diet may not inevitably depend on the availability of basic necessities. Further analysis (S1 table) revealed that rural participants who were in the category of high lived poverty presented reduced odds of having adequate dietary diversity (OR= 0.7, p = 0.476, 95%CI 0.3-1.7) while increased odds was observed among urban girls (OR=1.5, p = 0.301, 95%CI= 0.7-3.3) though the relationships were not significant. This indicates that the poorer participants in the rural areas did not have adequate dietary diversity which was evident in more than half of their population being classified in the high LPI group. However, the urban poor may consume nutritious diets as these are readily available in urban centers, thus, the little available food could be inexpensive, nutritious and diverse. Another reason could be the dependence of the urban on fast foods which are usually energy dense and monotonous.

Prevalence of food cravings in this study (64%) was similar to findings conducted in Ghana (52), Nigeria (53) and Iran (54) which were 67.7%, 61.3% and 60% respectively but was slightly lower than findings from Kenya (55) and Ecuador (56) which were 74% and 69% respectively. Food aversion prevalence (45%) was comparable to findings which were also conducted in Ghana (44.8%) by (52) and in Kenya (49%) by (57) but lower than those reported by (18) and (56) in Tanzania (70%) and Ecuador (74%) respectively. Current findings show practice of pica during pregnancy (39%) was slightly lower than studies conducted in Ghana by (46,52,58) which were between 47 – 48% but higher than those conducted in Kenya (57) and Ethiopia (59) which were 27% and 30% respectively. Regression analysis showed that participants who did not practice pica and food aversion had higher odds for adequate DD. However, (60,61) give credence to the fact that pica is commonly practiced among the poor communities than otherwise. This finding may imply that the practice of pica and food aversion could lead to reduced dietary intake and consequently micronutrient inadequacy.

### LIMITATION

The use of the FAO’s guidelines in measuring dietary diversity only takes into account the past 24-hour meal consumed by participants which might not reflect usual consumption habit. Also, it does not include the quantity of food items consumed as it only reflects economic access to food and not nutritional quality. There is no established cut off values for adequate dietary diversity as well. Also, dietary intake can vary during lean or bumper seasons as well as during festivities which were not considered during this study. Besides the limitations, this study has provided a new insight on factors that influence dietary diversity among pregnant adolescents to the scientific community especially in the Ghanaian context which is novel and interesting.

## 5.0 CONCLUSIONS

Dietary diversity among pregnant adolescents in the middle belt of Ghana is considered moderate although over half of these adolescents had low DD. The consumption of less diversified foods was higher among the rural pregnant adolescents and those aged 16-19 years. Being urban dweller, having higher income and practising good eating behaviours during pregnancy were significant determinants for adequate dietary diversity. Therefore, interventions to support and improve socio-economic and livelihoods of pregnant adolescents are likely to improve dietary diversity, and reduce the risk of malnutrition during pregnancy and adverse birth outcomes.

## Data Availability

Data cannot be shared publicly because it contains sensitive identifying information.
However, data are available from the Committee on Human Research Publication and
Ethics (CHRPE), the ethics board of the School of Medical Sciences of the Kwame
Nkrumah University of Science and technology, KNUST and Komfo Anokye Teaching
Hospital (KATH), Kumasi, Ghana for
researchers who meet the criteria for access to confidential data.

## DATA AVAILABILITY

The data that support the findings of this study are available from the corresponding author upon reasonable request.

## ACKNOWLEDGEMENT

We thank the directors of health and health workers in the health centers where the study took place, and most of all the pregnant adolescents who participated in the study.

## Supporting Information

**S1 TABLE.** PREDICTORS OF ADEQUATE DIETRY DIVERSITY AMONG COMMUNITY TYPES.

| Variables | $\beta$ | Significance | OR (95% CI) |
| --- | --- | --- | --- |
| **Rural** |  |  |  |
| Low Moderate |  |  | 1 |
| High Moderate | -0.832 | 0.040 | 0.4 (0.197 – 0.962) |
| High | -0.304 | 0.476 | 0.7 (0.320 – 1.701) |
| **Urban** |  |  |  |
| Low Moderate |  |  | 1 |
| High Moderate | -0.010 | 0.974 | 1.0 (0.544 - 1.804) |
| High | 0.414 | 0.301 | 1.5 (0.690 - 3.317) |

## REFERENCES

1. World Health Organisation. Preventing Early Pregnancy and Poor Reproductive Outcomes among adolescents in developing countries. WHO Guidelines. Geneva; 2011.

2. Nguyen PH, Sanghvi T, Tran LM, Afsana K, Mahmud Z, Aktar B, et al. The nutrition and health risks faced by pregnant adolescents: Insights from a cross-sectional study in Bangladesh. Ciccozzi M, editor. PLoS One [Internet]. 2017 Jun 8 [cited 2018 Sep 12]; 12(6):e0178878. Available from: http://dx.plos.org/10.1371/journal.pone.0178878

3. McGuire, Shelley. FAO, IFAD, and WFP. The State of Food Insecurity in the World 2015: Meeting the 2015 International Hunger Targets: Taking Stock of Uneven Progress. Rome: FAO, 2015. Adv Nutr. 2015;6(5):623-4.

4. Johnston, B. R. Transforming our world: the 2030 Agenda for sustainable development. Arsenic Research and Global Sustainability – Proceedings of the 6th International Congress on Arsenic in the Environment, AS 2016. 2016.

5. Lee SE, Talegawkar SA, Merialdi M CE. Dietary intakes of women during pregnancy in low- and middle-income countries. Public Heal Nutr. 2013; 16(08): 1350-3.

6. Desta, Melaku, Akibu M, Tadese M, Tesfaye M. Dietary Diversity and Associated Factors among Pregnant Women Attending Antenatal Clinic in Shashemane, Oromia, Central Ethiopia: A Cross-Sectional Study. J Nutr Metab [Internet]. 2019 Mar 12 [cited 2019 Oct 8];2019:1-7. Available from: https://www.hindawi.com/journals/jnme/2019/3916864/

7. Kennedy G, Ballard T, Dop M. Guidelines for measuring household and individual dietary diversity. FAO. 2010.

8. Onyango, A., Koski, K. G. & Tucker KL. Food diversity versus breastfeeding choice in determining anthropometric status in rural Kenyan toddlers. Int J Epidemiol. 1998;27:484-9.

9. Tarini, A., Bakari, S. & Delisle H. The overall nutritional quality of the diet is reflected in the growth of Nigerian children. Sante. 1999;16:23-31.

10. Yeneabat, T., Adugna, H., Asmamaw T et al. Maternal dietary diversity and micronutrient adequacy during pregnancy and related factors in East Gojjam Zone, Northwest Ethiopia, 2016. BMC Pregnancy Childbirth. 2019;(19): 173.

11. Archan Mukheijee, Sourabh Paull, Indranil Saha, Tapas Kumar Som GG. Dietary Diversity and its Determinants: A Community-Based Study among Adult Population of Durgapur, West Bengal. 2018;296-301.

12. Kiboi W, Kimiywe J, Chege P. Determinants of dietary diversity among pregnant women in Laikipia County, Kenya: a cross-sectional study. BMC Nutr. 2017;

13. Ballard, Terri, Deitchler JCASM. Household Hunger Scale: Indicator Definition and. Nutrition. 2011;(August).

14. Mattes, Robert. Afrobarometer; The material and political bases of lived poverty in Africa: insights from the Afrobarometer. 2008.

15. Evans, W. G, Kantrowitz E. Socioeconomic Status and Health: The Potential Role of Environmental Risk Exposure. Annu Rev Public Health. 2002;23(1):303-31.

16. Mowen AJ. Parks, Playgrounds and Active Living. 2010; 1-15.

17. Bell, J., Mora, G., Hagan, E., Rubin, V., & Karpyn A (2013). Access to Healthy Food and Why It Matters: A Review of the Research. 2013;368. Available from: http://www.policylink.org/find-resources/library/access-to-healthy-food-and-why-it-matters

18. Nyaruhucha, M CN. Food cravings, aversions and pica among pregnant women in Dar es Salaam, Tanzania. Tanzan J Health Res [Internet]. 2009 Jan;11(1):29-34. Available from: https://www.ncbi.nlm.nih.gov/pubmed/?term=Food+cravings%2C+aversions+and+pica+among+pregnant+women+in+Dar+es+Salaam%2C+Tanzania.+Tanzania+Journal+of+Health+Research

19. Orloff, C N, Hormes JM. Pickles and ice cream! Food cravings in pregnancy: hypotheses, preliminary evidence, and directions for future research. Front Psychol. 2014;5:1076.

20. Mulder, H EJ, Robles de Medina PG, Huizink AC, Van den Bergh BRH, Buitelaar JK, et al. Prenatal maternal stress: effects on pregnancy and the (unborn) child. Early Hum Dev. 2002 Dec;70(1-2):3-14.

21. Hainutdzinava N. Food Cravings and Aversions during Pregnancy: A Current Snapshot. J Pediatr Mother Care. 2017;02(01):1-5.

22. Meule, Adrian. Food craving: an overview. Die Ernährung. 2016 Jan 1;40:36-40.

23. Demissie T, Muroki NM, Kogi-Makau W. Food aversions and cravings during pregnancy: prevalence and significance for maternal nutrition in Ethiopia. Food Nutr Bull. 1998; 19(1):20–6.

24. Placek CD, Hagen EH. Fetal Protection: The Roles of Social Learning and Innate Food Aversions in South India. Hum Nat. 2015 Sep;26(3):255-76.

25. Young, M.F., Pressman E, Foehr ML, McNanley T, Cooper E, Guillet R, et al. Impact of maternal and neonatal iron status on placental transferrin receptor expression in pregnant adolescents. Placenta [Internet]. 2010 Nov [cited 2019 Mar 22];31(11): 1010-4. Available from: https://linkinghub.elsevier.com/retrieve/pii/S0143400410003140

26. Young, L. S, Khalfan SS, Farag TH, Kavle JA, Ali SM, et al. Association of pica with anemia and gastrointestinal distress among pregnant women in Zanzibar, Tanzania. Am J Trop Med Hyg. 2010;83(1): 144-51.

27. Awusabo-asare K, Biddlecom A, Kumi-Kyereme A, Patterson K. Adolescent Sexual and Reproductive Health in Ghana: Results from the 2004 National Survey of Adolescents [Internet]. Occasional Report No 22. 2006. 1-148 p. Available from: www.guttmacher.org.

28. University of Ghana, GroundWork, University of Wisconsin-Madison, KEMRI-Wellcome Trust, UNICEF. Ghana Micronutrient Survey 2017. Accra G 2017. Ghana Micronutrient Survey 2017. 2017.

29. Ghana Statistical Service. Population by region [Internet]. 2014 [cited 2019 Oct 30]. Available from: http://www.statsghana.gov.gh/regionalpopulation.php?population=MTI5MzE3OTU5OC40NDg1&&Ashanti&regid=1

30. Ghana Statistical Service (GSS). 2010 Population & Housing Census National Analytical Report. Ghana Stat Serv [Internet]. 2013;1-91. Available from: http://www.statsghana.gov.gh/gssmain/fileUpload/pressrelease/2010_PHC_National_Analytical_Report.pdf%0Ahttp://statsghana.gov.gh/docfiles/2010phc/National_Analytical_Report.pdf

31. Osman S, Saaka M, Siassi F, Qorbani M, Yavari P, Danquah I, et al. A comparison of pregnancy outcomes in Ghanaian women with varying dietary diversity: A prospective cohort study protocol. BMJ Open. 2016 Sep 1;6:e011498.

32. Quansah DY, Boateng D. Maternal dietary diversity and pattern during pregnancy is associated with low infant birth weight in the Cape Coast metropolitan hospital, Ghana: A hospital based cross-sectional study. Heliyon [Internet]. 2020 May 8;6(5):e03923-e03923. Available from: https://pubmed.ncbi.nlm.nih.gov/32420489

33. Ayensu J, Edusei A, Oduro I, Larbie C. Status of Some Antioxidant Micronutrient and Pregnancy Outcomes in Ghanaian Status of Some Antioxidant Micronutrient and Pregnancy Outcomes in Ghanaian Adolescents Attending Antenatal Clinic in Urban (Suntreso) and Rural (Mampong) Hospitals. EJNFS. 2017;7(2)(January):120-7.

34. FAO and FHI. Minimum Dietary Diversity for Women A Guide to Measurement. 2016.

35. Ampiah MKM, Kovey JJ, Apprey C, Annan RA. Comparative analysis of trends and determinants of anaemia between adult and teenage pregnant women in two rural districts of Ghana. BMC Public Health [Internet]. 2019; 19(1): 1379. Available from: https://doi.org/10.1186/s12889-019-7603-6

36. Ayamba JA. Relationship between dietary diversity and hemoglobin concentration among women in three communities in the Binduri District of the Upper East Region of Ghana [Internet]. University of Ghana. 2018. Available from: http://ugspace.ug.edu.gh

37. Ayensu J, Annan R, Lutterodt H, Edusei A. Prevalence of anaemia and low intake of dietary nutrients in pregnant women Prevalence of anaemia and low intake of dietary nutrients in pregnant women living in rural and urban areas in the Ashanti region of Ghana. PLoS One [Internet]. 2020;15(1)(January). Available from: http://dx.doi.org/10.1371/journal.pone.0226026

38. Nguyen PH, Huybregts L, Sanghvi TG, Tran LM, Frongillo EA, Menon P, et al. Dietary diversity predicts the adequacy of micronutrient intake in pregnant adolescent girls and women in Bangladesh, but use of the 5-group Cutoff Poorly identifies individuals with inadequate intake. J Nutr. 2018;148(5):790-7.

39. Abdirahman M, Chege P, Kobia J. Nutrition Knowledge and Dietary Practices among Pregnant Adolescents in Mandera County, Kenya. Food Sci Nutr Res. 2019;2(2): 1-8.

40. Beatriz M, Castro T De, Amélia A, Vilela F, Silva A, Oliveira D De, et al. Sociodemographic characteristics determine dietary pattern adherence during pregnancy Sociodemographic characteristics determine dietary pattern adherence during pregnancy. 2015;(January 2017).

41. Krieger J, Pestoni G, Cabaset S, Brombach C, Sych J, Schader C, et al. Dietary Patterns and Their Sociodemographic and Lifestyle Determinants in Switzerland: Results from the National Nutrition Survey menuCH. Nutrients. 2019;11(62).

42. Sintayehu H, Bedasa W. Dietary diversity and associated factors among pregnant women attending antenatal care at public health facilities in Bale Zone, Southeast Ethiopia. 2019;

43. Bagson, And E, Naamwintome Beyuo A. Home Gardening: The Surviving Food Security Strategy in the Nandom Traditional Area-Upper West Region Ghana. J Sustain Dev Africa. 2012; 14(1).

44. Ecker O, Tan TJ-Fr, Alpuerto V, Diao X. Economic Growth and Agricultural Diversification Matters for Food and Nutrition Security in Ghana. Transfroming Agric Conf. 2012;(November): 1-5.

45. Barnett, I, Srivastava S. External evaluation of mobile phone technology-based nutrition and agriculture advisory services in Africa and Desk-review: Smallholder farming, nutrition and m-Agriculture services in Ghana. 2017.

46. Mensah, O. F, Twumasi P, Amenawonyo XK, Larbie C, Jnr AKB. Pica practice among pregnant women in the Kumasi metropolis of Ghana. Int Health [Internet]. 2010;2(4):282-6. Available from: http://dx.doi.org/10.1016/j.inhe.2010.09.004

47. Ruel, T. M, Deitchler M, Arimond M. Developing Simple Measures of Women’s Diet Quality in Developing Countries: Overview. J Nutr. 2010;140(11):2048S-2050S.

48. Ochieng J, Victor A.S, Lukumay P. J and DT. “Determinants of dietary diversity and the potential role of men in improving household nutrition in Tanzania,.” PLoS One. 2017; 12(12).

49. Bailey, L R, Akabas SR, Paxson EE, Thuppal S V, Saklani S, et al. Total Usual Intake of Shortfall Nutrients Varies With Poverty Among US Adults. J Nutr Educ Behav. 2017 Sep;49(8):639-646.e3.

50. Odhiambo FO. Assessing the Predictors of Lived Poverty in Kenya: A Secondary Analysis of the Afrobarometer Survey 2016. J Asian Afr Stud. 2019;54(3):452-64.

51. Habyarimana F, Zewotir T, Ramroop S. Determinants of Poverty of Households in Rwanda: An Application of Quantile Regression. J Hum Ecol. 2015 Jun 1;50:19-30.

52. Koryo-Dabrah A, Nti CA, Adanu R. Detary practices and nutrient intakes of pregnant women in Accra, Ghana. Curr Res J Biol Sci [Internet]. 2012;4(4):358-65. Available from: http://maxwellsci.com/print/crjbs/v4-358-365.pdf

53. Olusanya JO, Ogundipe FO. Food aversion and craving among pregnant women in Akure, Ondo state, Nigeria. Int J Trop Med. 2009 Jan 1;4:100-3.

54. Khoushabi, Fahimeh, Ahmadi P, Shadan MR, Heydari A. Pica Practices among Pregnant Women Are Associated with Lower Hemoglobin Levels and Pregnancy Outcome. 2014;(August):646-52.

55. Kariuki L, Lambert C, Purwestri R, Konrad H. Trends and consequences of consumption of food and non-food items (pica) by pregnant women in Western Kenya. NFS [Internet]. 2016;5(January 2015): 1-4. Available from: http://dx.doi.org/10.1016/j.nfs.2016.09.001

56. Weigel, Margaret, Coe K, Castro N, Caiza M, Tello N, et al. Food Aversions and Cravings during Early Pregnancy: Association with Nausea and Vomiting Food Aversions and Cravings during Early Pregnancy: Association with Nausea and Vomiting. 2011;(June 2014).

57. Kariuki L, Lambert C, Purwestri R, Biesalski HK. Trends and consequences of consumption of food and non-food items (pica) by pregnant women in Western Kenya. NFS J [Internet]. 2016;5(November 2014):1-4. Available from: http://dx.doi.org/10.1016/j.nfs.2016.09.001

58. Konlan K, Abdulai J, Konlan K, Amoah R, Doat AR. Practices of pica among pregnant women in a tertiary healthcare facility in Ghana. Nurs Open. 2020 Jan 28;7.

59. Yoseph, Handiso H. Prevalence of food aversions, cravings and pica during pregnancy and their association with nutritional status of pregnant women in Dale Woreda, Sidama zone, SNNPRS, Ethiopia. Int J Nutr Metab. 2015;7(1): 1-14.

60. Knox, B, Kremer J PJ. A survey of dietary urges and consumption during pregnancy in Belfast working class women. 1995;1:125-44. Soc Sci Heal. 1995;(1): 125-44.

61. Lacey, E. P. Broadening the perspective of pica: Literature review. Public Health Rep. 1990; 105(1):29-35.Supporting Information

